# Behavioral Responses to Fevers and Other Medical Events in Children With and Without ASD

**DOI:** 10.1101/2022.05.23.22275374

**Authors:** Katherine Byrne, Shuting Zheng, Somer Bishop, Juliana Boucher, Sheila Ghods, So Hyun Kim, Catherine Lord

**Author notes:** Address correspondence to: Katherine Byrne, Semel Institute for Neuroscience and Human Behavior, 760 Westwood Plaza 68-265, Los Angeles, CA 90024, [ ]. **Role of Funder/Sponsor:** The Simons Foundation had no role in the design and conduct of the study. **Clinical Trial Registration:** N/A. **Contributors’ Statement Page** Katherine Byrne collected data, drafted the initial manuscript, and reviewed and revised the manuscript. Dr. Zheng carried out the initial analyses and reviewed and revised the manuscript. Drs. Bishop, Kim, and Lord designed the data collection instrument, conceptualized and designed the study, supervised data collection, and reviewed and revised the manuscript. Juliana Boucher designed the data collection instrument, collected data, and reviewed and revised the manuscript. Sheila Ghods collected data and reviewed and revised the manuscript. All authors approved the final manuscript as submitted and agree to be accountable for all aspects of the work.

## Abstract

**Objective:** Anecdotal reports and a small number of research studies suggest possible behavioral improvements in children with autism spectrum disorders (ASD) during a fever. However, previous studies rely largely on retrospective reports of this phenomenon. Establishing a robust association between fever and reduction of ASD-related symptoms would promote opportunities for the development of innovative therapeutic interventions for children with ASD.

**Methods:** Prospective data were collected from 141 children with ASD and 103 typically developing (TD) controls using parent responses to an 11-item behavioral survey. Behaviors when no illness was present, during a fever, the week after a fever, and during non-febrile illnesses for TD and ASD children were compared. Profiles of cases in which caregivers reported consistent behavioral improvements during fever are described.

**Results:** Data indicated worsening social, emotional/behavioral, and somatic symptoms during a fever regardless of diagnosis, with children with ASD demonstrating greater worsening of behaviors during a fever than TD children. Three children (all with ASD) out of 244 demonstrated consistent behavioral improvements during a fever; these children had a range of cognitive and adaptive skills.

**Conclusions:** Children with ASD had stronger negative responses to fever than TD children. These findings contradict previous literature suggesting behavioral improvements for children with ASD. While improvements may occur for some children, it does not appear to be a common phenomenon. Additional research is needed to elucidate the nature of behavioral improvements in the subset of children with ASD who may respond positively to fever.

**Article Summary:** This study examines behavioral changes during fever and other medical events in children with autism compared to behavioral changes in a typically developing control group.

**What’s Known on this Subject:** Previous research and consistent anecdotal reports suggest the possibility of behavioral improvements for children with autism during a fever. There is a dearth of prospectively collected data examining these effects.

**What This Study Adds:** Children with autism consistently had stronger and more frequent negative behavior changes during fever than typical children who also primarily showed worsening of behavior during fevers. Three autistic children, and no typical children, showed improvements in varied areas during fevers.

---

Anecdotal reports suggesting the possibility of behavioral improvements in children with autism when they experience a fever have been made for years.^1^ Yet, there is surprisingly little published data examining these effects. The largest study to date used retrospective caregiver-report data from a sample of over 2000 children with autism spectrum disorder (ASD) from the Simons Simplex Collection (SSC). Seventeen percent of families reported improvements in autism symptoms during a fever.^2^ This subgroup had less speech, lower nonverbal IQs, higher levels of autism symptoms, higher levels of restricted and repetitive behaviors, and higher parent-reported levels of stereotypies, inappropriate speech, hyperactivity, and irritability as measured on the Aberrant Behavior Checklist (ABC)^3^, compared to children whose caregivers did not report behavioral improvements. However, this study relied solely on retrospective parent-report and was collected in response to just one (arguably leading) question: “Does your child seem to show any improvement in symptoms of autism when he/she has a fever?”^3^ which created the potential for recall bias and inaccurate reporting.

In another study, Curran et al^4^ prospectively compared scores on the ABC during a fever, after a fever, and during no fever. Eighty-three percent of caregivers reported fewer aberrant behaviors on at least one domain of the ABC during a fever. Improvements on the “lethargy/social withdrawal” subscale did not account for the behavioral changes. While use of prospective report is a significant strength of this study, data were derived from a very small sample of children with ASD who developed a fever (*n* = 30) with no comparison group of children with nonfebrile illnesses. Thus, no data currently exist comparing behavior during fevers vs. non-febrile illnesses in children with ASD, or in comparison to children without ASD. Additionally, while families were blind to the researchers’ hypotheses, they, nonetheless, were aware that this study was testing behavioral responses specifically to fevers, which could increase the possibility of biased reporting.

The current study aimed to investigate, prospectively, the association between autism and behavioral improvements during fever and non-febrile medical events in children with and without ASD. Parent reports were collected at baseline (i.e., no illness), during fevers, and during non-febrile illnesses in a sample of 141 children with ASD and 103 TD controls. We set out to understand the degree to which children with ASD have unique responses to fevers by asking the following questions: (1) Do participants exhibit behavioral changes during a fever? If so, does the nature of these changes differ for children diagnosed with ASD? (2) Do behavioral changes during a fever differ from behavioral changes during non-febrile illnesses? (3) What are the phenotypic characteristics of children who exhibit positive behavioral changes during a fever?

## Methods

### Participants

This study included 244 children; 141 were diagnosed with ASD, and 103 were TD. The majority of participants (70%) were male. The parent-reported racial/ethnic breakdown of the participants is as follows: 55% White, 16.4% Asian, 5.3% Black, 17.6% Mixed Race, and 5.7% American Indian, Pacific Islander, or Other; 24.6% identified as being Hispanic. Table 1 presents the demographic characteristics of those who had a fever and those who did not, by diagnosis.

**Table 1.** Demographic Differences by Fever and Diagnosis

|  | <u>Fever</u> |  | <u>No Fever</u> |  |
| --- | --- | --- | --- | --- |
|  | ASD<br>(n = 50) | No ASD<br>(n = 43) | ASD<br>(n = 91) | No ASD<br>(n = 60) |
| Demographic Characteristics |  |  |  |  |
| Age at intake<br>Mean (SD) | 5.06 (1.6) ** | 3.37 (1.4) ** | 4.7 (1.6) | 4.2 (1.6) |
| Gender (Males)** | 80%** | 51%** | 82%** | 56%** |
| Race (Non-White) | 42% | 39% | 49% | 45% |
| Ethnicity (Hispanic) | 24% | 16% | 31% | 20% |
| Behavioral Measures<br>Mean (SD) |  |  |  |  |
| PPVT **<br>Standard Score | 92.75 (23.1) | 115.27 (11.4) | 91.38 (23.2) | 116.56 (14.8) |
| PSI **<br>Total Stress Score | 105.59 (18.9) | 69.88 (20.1) | 101.13 (21.5) | 67.93 (16.2) |
| ABC – Irritability **<br>(intake) | 12.15 (9.1) | 3.39 (4.1) | 11.72 (8.4) | 3.27 (5.9) |
| ABC – Hyperactivity **<br>(intake) | 18.16 (10.2) | 4.37 (6.4) | 18.13 (11.1) | 4.02 (7.5) |
| Vineland ABC **<br>Standard Score | 70.27 (15.3) | 101.12 (10.6) | 73.5 (16.2) | 101.1 (13.3) |
| SRS **<br>Total T-Score | 76.43 (11.0) | 45.0 (9.3) | 71.08 (11.56) | 46.08 (7.1) |
| CBCL (1.5-5) Internalizing **<br>T-score | 62.04 (10.3) | 41.58 (14.6) | 59.16 (15.9) | 42.61 (10.8) |
| CBCL (1.5-5) Externalizing **<br>T-score | 57.98 (11.3) | 41.81 (14.4) | 56.19 (16.0) | 42.26 (12.4) |

Note: \* $p < .05$ ; \*\* $p < .01$
Fever group is defined by the individuals who had at least one fever $\geq 100.0$ degrees Fahrenheit

Children were recruited through research databases, word-of-mouth referrals, and flyers posted in local autism clinics, pediatric offices, daycares, and schools in the greater Los Angeles, San Francisco, and New York areas. Inclusion criteria included being between the ages of 2 to 7 years, having had a fever in the previous 12 months, and independently walking. If recruited for the ASD cohort, parents were asked to provide documentation of a clinical ASD diagnosis for the child. Exclusion criteria included severe sensory impairment, or, if recruited for the TD cohort, a diagnosis or receipt of services for a developmental or psychiatric disorder.

### Procedures

Event reporting and behavioral tracking data were collected through a smartphone application developed for the current study. During the initial study visit, research staff provided the participating parent with instructions on how to download and operate the application used for data collection. Parents and children completed a battery of questionnaires, and assessments described below.

Behavioral tracking lasted for three months. Once per week (at a time when no event had occurred), parents were asked to complete the behavioral survey; these data were collected as baseline measures of behavior. Parents also used the app to report on medical events, as well as other “out of the ordinary” life events that occurred, such as non-routine vacations. If parents reported an event, they were asked to explain the event that transpired and complete the behavioral survey (see Supplement 1). Each time parents submitted an event and/or completed the behavioral survey, they were asked to take and report their child’s temperature. Non-medical data were collected to conceal the specific aims and hypotheses of the study (pertaining to behavioral responses to fever) and prevent biased reporting. To address the aims of the current study, only data regarding medical events were analyzed.

After the three-month data collection period concluded, participants completed a subset of questionnaires. Compensation was provided to all participating families. Researchers acquired informed consent from parents with approval from the University of California, Los Angeles (UCLA), University of California San Francisco (UCSF), and Weill Cornell Medical College Institutional Review Boards.

### Measures

#### Event and Behavioral Survey Reporting via App

All event data were coded into one of five categories: baseline, fever, week-after-fever, non-fever medical, and other. Baseline data are those that parents submitted once per week at a time when no event had occurred. Fever data are defined as medical events in which the parent reported the child to have a fever ≥ 100.0 degrees Fahrenheit. Week-after-fever data are defined as data that were reported in the 7 days succeeding the final fever report. These data were considered separately from baseline to examine whether the effects of fever may last beyond the fever itself and to prevent confounding the baseline data. Non-fever medical data are defined as any medical event (e.g., gastrointestinal issues, sore throat) that occurred without a coinciding fever. “Other” data are defined as various “out of the ordinary” events that were not medical in nature.

The behavioral survey, created by the study team, was comprised of 11 questions regarding the child’s mood and behavior. The survey asked parents to rate, on a five-point Likert scale, how much the child exhibited a certain behavior compared to usual behavior. Lower scores indicated less than usual and higher scores indicated more than usual. For data analysis, questions nine and ten (i.e., irritability and anxiety) were reverse coded to ensure that scores of five indicated lessening behavioral symptoms (i.e., less irritability and less anxiety). For ease of interpretability, all items were then recoded to scores from -2 to 2; negative scores indicated worsening symptoms, and 0 represented the same as usual.

#### Other Measures

We collected various measures to characterize the sample and examine individual differences in behavioral responses to fever. The PPVT-4^5^ is an assessment of receptive vocabulary for individuals between the ages of 2 years, 6 months through adulthood. If a child was unable to obtain a basal score on the PPVT-4, the nonverbal subtests (Visual Reception and Fine Motor) of the Mullen Scales of Early Learning (MSEL)^6^ were administered. These tests were administered to provide an estimate of cognitive ability. Standard scores from the PPVT were used, whereas age equivalents from the MSEL were used to calculate a ratio IQ/developmental quotient.^7^ The Vineland-3^8^ is a semi-structured parent interview that assesses adaptive functioning in individuals from birth through adulthood. Scores from the overall Adaptive Behavior Composite (ABC) were used to provide an estimate of adaptive functioning for each participant. The SRS-2^9^ is a parent-rated questionnaire that was collected to measure the severity of ASD-related symptoms. The PSI-4 SF^10^ is a parent-rated questionnaire that was collected to assess parents’ level of stress in the parent-child relationship. The ABC-C^3^ is a parent-rated questionnaire that measures the behavior problems of individuals with ASD and other developmental disabilities. For purposes of this study, we used the Irritability and Hyperactivity subscales. Finally, the CBCL/1.5-5 and CBCL/6-18^11^ are parent-rated questionnaires that assess the emotional and behavioral problems of children. For purposes of this study, we used the Internalizing and Externalizing subscales.

### Statistical Analyses

#### Preliminary Analyses

##### Factor Analyses of Behavioral Survey

Exploratory factor analysis (EFA) with oblique rotation was employed to generate the latent constructs measured by the behavioral survey. EFA demonstrated that a three-factor structure fit best for the survey items: Factor 1 (“Social Behaviors”) consisted of six items: happy/cheerful, talk to others, physically active, social/interested, communicating ideas, and play; Factor 2 (“Emotional and Behavioral Problems”) consisted of three items: anxious, managing behaviors, and irritable; Factor 3 (“Somatic Symptoms”) consisted of two items: eat and sleep (see Supplement 2 for detailed results). The average rating of the items within each factor was calculated to represent factor scores because the number of items loading on each factor differed.

##### Sample Characteristics

Descriptive statistics regarding demographic and behavioral characteristics of the sample were generated. Independent samples t-tests were used to compare these characteristics between the ASD and TD cohorts (see Table 1).

##### Multilevel Modeling

Parent-report behavioral ratings across multiple days nested within participants were used. Multilevel models with behavioral rating factor scores as outcomes were used to account for the nesting and to capture both fixed and random effects at two levels of analysis. Level 1 focused on within-individual effects; we tested whether parents were more likely to report children’s behaviors to deviate (positively or negatively) from what they usually exhibited on days when they experienced a fever, and the week after a fever, as compared to behaviors at baseline. Level 2 focused on between-individual effects; we tested whether the within-person association between behavioral changes and fever conditions differed for the ASD vs. TD samples. Sequential multilevel models were specified in SAS PROC Mixed to address the research questions respectively. Model fit indices (i.e., AIC, BIC, smaller values mean better model fit), variances explained, and parameter estimates of the fixed and random effects in each model are reported.

To address research questions of behavioral changes across fever conditions: First, the intraclass correlation (ICC) of each factor score was computed to reflect how much of the total variation in behavioral ratings was accounted for by individuals through the unconditional model (Model 1) with no predictors. Second, to test the effect of fever conditions, we estimated separate two-level models with each of the three factor scores as the outcome variable. We included the fixed effect of the fever condition variable (three categories: baseline, fever, and week-after) with baseline as the reference condition at Level 2 (individual level), and the random intercept at Level 1 to capture random between-individual variability in children’s average level of behavioral changes on the factors and the random slope of between-individual variability in associations between fever conditions and behavioral changes (Model 2). Third, we added the fixed effect of diagnosis (with TD as the reference) and the interaction effect of diagnosis and fever condition to Model 2 (Model 3). Model 3 estimates were used to determine the main effect of diagnosis on behavior rating factor scores and to examine whether children with ASD were affected differently by fever conditions than TD children. Lastly, multi-level models were fitted to test the behavioral changes during fever compared to during non-febrile illnesses, and whether these changes differed by diagnosis.

##### Case Analysis of Children with Behavioral Improvement During Fever

We describe profiles of cases in which caregivers reported behavioral improvements (score of 5 indicating clear changes) on the same behavioral survey item(s) across at least two separately reported fevers.

## Results

### Descriptive Analyses of Behavioral Ratings

Table 2 presents the average number of baseline, fever, week-after-fever, and non-febrile illnesses reported for children, by diagnosis. On average, parents provided 8 baseline survey reports, plus 2 during a fever, 1.5 the week after a fever, and 2.5 during non-febrile medical illnesses. A similar proportion of parents of children with and without ASD (50 and 41, respectively) reported at least one fever event (*M*_*FeverReports*_ = 2.04); fewer parents completed behavioral survey ratings in the week after a fever compared to all other conditions.

**Table 2.** Number of Behavioral Survey Ratings by Diagnosis

|  | Baseline |  |  |  | Fever |  |  |  | Week After Fever |  |  |  | Non-Fever Medical Event |  |  |  |
| --- | --- | --- | --- | --- | --- | --- | --- | --- | --- | --- | --- | --- | --- | --- | --- | --- |
|  | N | Mean | SD | Range | N | Mean | SD | Range | N | Mean | SD | Range | N | Mean | SD | Range |
| ASD | 134 | 8.25 | 4.68 | [1, 31] | 50 | 1.98 | 1.38 | [1, 7] | 29 | 1.90 | 1.95 | [1, 11] | 98 | 2.65 | 2.10 | [1, 10] |
| Non-ASD | 102 | 8.23 | 3.57 | [1, 21] | 41 | 2.10 | 1.80 | [1, 11] | 16 | 1.38 | 0.81 | [1, 4] | 63 | 2.58 | 2.28 | [1, 13] |

### Behavioral Changes During Fever vs. Baseline

Multilevel modeling revealed a significant main fixed effect of fever condition on all three factors, suggesting that children’s social, emotional/behavioral, and somatic symptoms all worsened during a fever compared to baseline (see Table 3 for test statistics). When comparing behaviors during the week after a fever to baseline, the only significant difference was in emotional and behavioral problems (Factor 2; see Table 3). The random slopes of fever conditions on behavior changes across all three factors were significant, suggesting that fever conditions differed for all types of behaviors (see Table 3). The fixed and random effect of fever condition explained 67% of the variance in Factor 1 behavioral ratings, 36% in Factor 2, and 43% in Factor 3 (see Table 3 for pseudo R^2^ values).

**Table 3.** Multilevel Model Comparing Behavior Changes During Fever and the Week After to Baseline

|  |  | Factor 1 |  |  |  | Factor 2 |  |  |  | Factor 3 |  |  |  |
| --- | --- | --- | --- | --- | --- | --- | --- | --- | --- | --- | --- | --- | --- |
|  |  | Model 2 |  | Model 3 |  | Model 2 |  | Model 3 |  | Model 2 |  | Model 3 |  |
| **Fixed Effects** |  | **B (SE)** | **p** | **B (SE)** | **p** | **B (SE)** | **p** | **B(SE)** | **p** | **B(SE)** | **p** | **B(SE)** | **p** |
| *Condition* | Baseline | -- | -- | -- | -- | -- | -- | -- | -- | -- | -- | -- | -- |
|  | Fever | -0.72<br>(0.05) | < .001 | -0.58<br>(0.07) | < .001 | -0.19<br>(0.05) | < .001 | -0.20<br>(0.07) | 0.009 | -0.43<br>(0.05) | < .001 | -0.31<br>(0.07) | < .001 |
|  | Week After<br>Fever | -0.14<br>(0.07) | 0.057 | 0.00<br>(0.12) | 0.993 | -0.24<br>(0.07) | 0.001 | -0.03<br>(0.13) | 0.820 | -0.05<br>(0.07) | 0.455 | -0.01<br>(0.12) | 0.96 |
| *Diagnosis* | Typically<br>Developing | -- | -- | -- | -- | -- | -- | -- | -- | -- | -- | -- | -- |
|  | ASD | -- | -- | 0.02<br>(0.05) | .732 | -- | -- | 0.06<br>(0.04) | 0.187 | -- | -- | 0.07<br>(0.04) | 0.111 |
| *Interaction*<br>*Effects* | ASD * Fever | -- | -- | -0.26<br>(0.10) | 0.011 | -- | -- | 0.01<br>(0.10) | 0.898 | -- | -- | -0.22<br>(0.10) | 0.023 |
|  | ASD * Week<br>After | -- | -- | -0.22<br>(0.15) | .153 | -- | -- | -0.32<br>(0.16) | 0.043 | -- | -- | -0.08<br>(0.15) | 0.583 |
| **Random Effects**<br>**(covariances)** |  | **Estimate** | **p** | **Estimate** | **p** | **Estimate** | **p** | **Estimate** | **p** | **Estimate** | **p** | **Estimate** | **p** |
|  | Intercept | 0.24 (0.01) | < .001 | 0.02<br>(0.01) | < .001 | 0.04<br>(0.01) | < .001 | 0.04<br>(0.01) | < .001 | 0.04<br>(0.01) | < .001 | 0.04<br>(0.01) | < .001 |
|  | Condition | 0.07 (0.01) | < .001 | 0.08<br>(0.01) | < .001 | 0.04<br>(0.01) | < .001 | 0.04<br>(0.01) | < .001 | 0.04<br>(0.01) | < .001 | 0.04<br>(0.01) | < .001 |
| **Model Fit Indices** |  |  |  |  |  |  |  |  |  |  |  |  |  |
|  | AIC | 2871 |  | 2871 |  | 3606 |  | 3610 |  | 3258 |  | 3262 |  |
|  | BIC | 2881 |  | 2881 |  | 3617 |  | 3620 |  | 3269 |  | 3272 |  |
|  | Pseudo-R<sup>2</sup> | 0.67 |  | 0.66 |  | 0.36 |  | 0.35 |  | 0.43 |  | 0.43 |  |

There was no overall effect of ASD diagnosis for any of the behavioral rating factor scores. However, a significant interaction was observed for all three behavioral factor scores. Compared to TD children, children with ASD were reported to have worse social (Factor 1) and somatic (Factor 3) factor scores during fever than during baseline. Children with ASD also showed worse emotional/behavioral problems (Factor 2) during the week after a fever compared to baseline (*p*s<.05). However, no significant improvements in model fit were observed when the main effect of ASD diagnosis and the interaction effect were added.

### Behavioral Changes During Fever vs. Non-febrile Illness

Multilevel models comparing behavioral changes during a fever to non-febrile illnesses demonstrated that caregivers reported their children to have worse social and emotional/behavioral factor scores during fever than during non-febrile illnesses (see Table 4 for test statistics). The addition of the main fixed effect of diagnosis and the interaction effect of diagnosis and condition did not improve model fit for any of the behavioral rating factor scores.

**Table 4.** Multilevel Model Comparing Behavior Changes During Fever to Non-Fever Medical Events

|  |  | <u>Factor 1</u> |  |  |  | <u>Factor 2</u> |  |  |  | <u>Factor 3</u> |  |  |  |
| --- | --- | --- | --- | --- | --- | --- | --- | --- | --- | --- | --- | --- | --- |
|  |  | Model 2 |  | Model 3 |  | Model 2 |  | Model 3 |  | Model 2 |  | Model 3 |  |
| **Fixed Effects** |  | **B**<br>**(SE)** | ***p*** | **B (SE)** | ***p*** | **B (SE)** | ***p*** | **B(SE)** | ***p*** | **B(SE)** | ***p*** | **B(SE)** | ***p*** |
| *Condition* | Non-Fever Medical | -- | -- | -- | -- | -- | -- | -- | -- | -- | -- | -- | -- |
|  | Fever | -0.40<br>(0.05) | < .001 | -0.29<br>(0.08) | < .001 | 0.04<br>(0.06) | 0.531 | 0.01<br>(0.09) | 0.918 | -0.23<br>(0.06) | < .001 | -0.17<br>(0.09) | 0.075 |
| *Diagnosis* | Typically Developing | -- | -- | -- | -- | -- | -- | -- | -- | -- | -- | -- | -- |
|  | ASD | -- | -- | -0.05<br>(0.07) | <.001 | -- | -- | 0.01<br>(0.08) | 0.894 | -- | -- | -0.01<br>(0.08) | 0.878 |
| *Interaction Effect* | ASD * Fever | -- | -- | -0.20<br>(0.10) | 0.056 | -- | -- | 0.05<br>(0.12) | 0.681 | -- | -- | -0.11<br>(0.12) | 0.351 |
| **Random Effects**<br>**(covariances)** |  | **Estimate** | ***p*** | **Estimate** | ***p*** | **Estimate** | ***p*** | **Estimate** | ***p*** | **Estimate** | ***p*** | **Estimate** | ***p*** |
|  | Intercept | 0.04<br>(0.02) | 0.034 | 0.04<br>(0.02) | 0.021 | 0.08<br>(0.04) | 0.016 | 0.07<br>(0.04) | 0.018 | 0.10<br>(0.03) | < .001 | 0.10<br>(0.03) | < .001 |
|  | Condition | 0.06<br>(0.02) | 0.005 | 0.05<br>(0.02) | 0.014 | 0.05<br>(0.03) | 0.063 | 0.05<br>(0.03) | 0.058 | 0.00<br>(0.03) | 0.466 | 0.00<br>(0.03) | 0.435 |
| **Model Fit Indices** |  |  |  |  |  |  |  |  |  |  |  |  |  |
|  | AIC | 743 |  | 741 |  | 973 |  | 979 |  | 1089 |  | 1093 |  |
|  | BIC | 753 |  | 751 |  | 983 |  | 988 |  | 1099 |  | 1103 |  |
|  | Pseudo-R<sup>2</sup> | 0.56 |  | 0.52 |  | 0.36 |  | 0.37 |  | 0.09 |  | 0.11 |  |

### Cases with Behavioral Improvements During Fever

Figure 1 depicts the proportion of participants who reported positive, negative, and no changes in response to each behavioral survey item. Three participants, all with ASD between the ages of 2-7, showed consistent positive changes on the same behavioral survey items during at least two fevers. Two participants were reported to be less irritable and have behaviors that were easier to manage during two out of two or three fevers. The third participant was reported to be less anxious and less irritable during two out of two fevers. Table 5 depicts the SRS-2, Vineland-3, and PPVT-4 scores of theses participants.

**Table 5.** Case Reports of Participants with Consistent Improvement

|  | Participant #1 | Participant #2 | Participant #3 |
| --- | --- | --- | --- |
| Diagnosis | ASD | ASD | ASD |
| Behavioral Improvements | Irritability, Happiness, Bx Management | Irritability, Bx Management | Anxiety, Irritability |
| PPVT Standard Score | 93 | 75 | 117 |
| Vineland ABC Standard Score | 92 | 63 | 101 |
| SRS Total T-Score | 67 | 88 | 94 |

**Figure 1.**
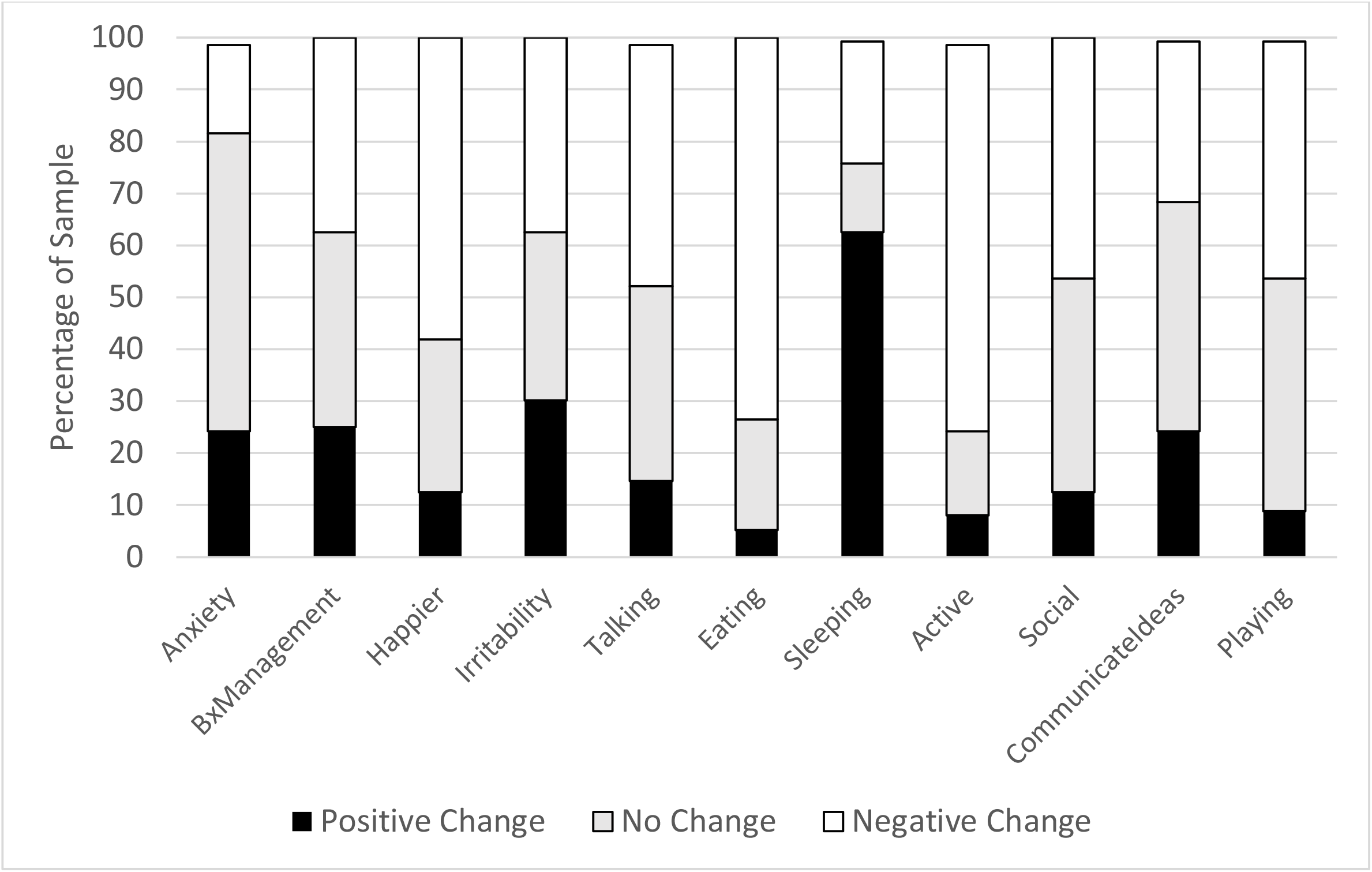
Proportion of participants who exhibited positive, negative, and no changes in behavior during a fever.

## Discussion

Results suggest that children with and without ASD demonstrate similar patterns of behavioral changes in response to fever. In general, children in both groups were reported to exhibit worse social, emotional/behavioral, and somatic symptoms, with these negative changes even more pronounced among children with ASD. These findings are opposite to numerous anecdotal reports and at least two empirical studies suggesting behavioral improvements during fever in children with autism.^4^ Nevertheless, our finding that three children with ASD demonstrated behavioral improvements in response to fever suggest that, while it is certainly not the norm, a small subset of children may indeed be positively affected by fever.

Previous data from the SSC reported that improvements associated with fever were more likely to occur in children with lower cognitive abilities and higher levels of restricted and repetitive behaviors. In contrast, many of the children in the current sample diagnosed with ASD had average or near-average cognitive abilities, which may help explain differences in fever responses in our sample. Yet, the three individuals who had consistent behavioral improvements with fevers varied in terms of cognitive and adaptive abilities. Their clear individual differences complicate potential conclusions about phenotypic patterns related to behavioral improvements during a fever for this sample.

It is possible that predictors of improvement are not at the phenotypic level but, rather, at the biological level. Some hypothesized mechanisms for improvement in autism symptoms during fever include the role of the locus coeruleus,^12^ inflammatory processes in the cerebellum,^13^ and hypothalamic responses to thermoregulation.^14^ Additional research could elucidate the nature of the association between fever and behavioral changes in children with ASD. If this phenomenon could be detected reliably in a subset of children with ASD, innovative therapeutic interventions could be developed from an understanding of the neuronal systems involved in symptom improvement as a response to fever. However, given the apparent rarity of this phenomenon, much larger sample sizes would be needed if researchers wanted to identify subgroups of individuals who respond positively to fever.

The ability to compare behavioral changes during a fever to during non-febrile illnesses was a noteworthy strength of this study’s methodology. Results demonstrated that most participants, TD or ASD, exhibited worsening social and emotional/behavioral symptoms during a fever compared to non-febrile illnesses. This suggests that it could be the fever itself or possibly the severity of the illness which we did not rate, as opposed to general immune responses, that are associated with behavioral changes.

This study had a number of limitations. Our sample size was relatively small, leading to limited power in analyses. Additionally, future studies could include a more diverse sample with children who are older and have varying cognitive abilities.

## Conclusions

This study demonstrated that both TD children and those with ASD display worsening social, emotional and behavioral, and somatic symptoms during fever, even when compared to non-febrile illnesses. Furthermore, children with ASD were more affected by fevers (i.e., greater worsening of symptoms) than TD children. Our findings suggests that, while behavioral improvements during fever may happen for a small subgroup of children with ASD, it likely is not the norm.

## Data Availability

All data produced in the present study are available upon reasonable request to the authors

## Abbreviations

(ASD): Autism spectrum disorder;
(TD): typically developing.

