## Supplement Table 1 Figure 1 for "Behavioral Responses to Fevers and Other Medical Events in Children With and Without ASD"

Supplement I: Behavioral Survey Questions

1. In the last 24 hours, how much has s/he been sleeping?

less than usual	same as usual	more than usual

2. In the last 24 hours, how much has s/he been eating?

less than usual	same as usual	more than usual

3. In the last 24 hours, how well has s/he played by her/himself or with others?

worse than usual	same as usual	better than usual

4. In the last 24 hours, how much did s/he talk to others?

less than usual	same as usual	more than usual

5. In the last 24 hours, how well could s/he communicate his/her ideas?

less than usual	same as usual	more than usual

6. In the last 24 hours, how social and interested in people was s/he?

less than usual	same as usual	more than usual

7. In the last 24 hours, how physically active has s/he been?

less than usual	same as usual	more than usual

8. In the last 24 hours, how easy was it to manage his/her behaviors?

harder than usual	same as usual	easier than usual

9. In the last 24 hours, how irritable has s/he been?

less than usual	same as usual	more than usual

10. In the last 24 hours, how anxious has s/he been?

less than usual	same as usual	more than usual

11. In the last 24 hours, how happy and cheerful has s/he been?

less than usual	same as usual	more than usual

Supplement II: Exploratory Factor Analysis of Behavioral Survey

Exploratory factor analysis with oblique rotation was employed to generate the latent constructs measured by the daily reports of behavioral ratings using SAS procedure PROC FACTOR. The optimal number of factors was determined by the number of factors with eigenvalues larger than 1 and the elbow point on the scree plot. Items were considered under the factor, where they have the largest factor loadings (absolute value) greater than 0.3.

Three factors were retained based on the eigenvalue criterion and Scree Plot examination (see Figure S2). When arriving at a simple factor structure (each item loads exclusively on one factor), the average rating of all the items was calculated to represent factor scores, respectively, considering the number of items loading on each factor might differ.

| Table S2 Factor Loadings of Behavioral Rating Items of the 3-Factor Structure | | | |
| --- | --- | --- | --- |
|  | Factor 1:  Social Behaviors | Factor 2:  Emotional and Behavioral Problems | Factor 3:  Somatic Symptoms |
| Q5: Talk to Others | ***0.87*** | -0.09 | -0.07 |
| Q9: Social/Interested | ***0.84*** | 0.00 | 0.00 |
| Q8: Physically Active | ***0.69*** | -0.04 | -0.07 |
| Q10: Communicate Ideas | ***0.66*** | -0.04 | 0.04 |
| Q11: Play | ***0.65*** | 0.21 | 0.03 |
| Q3: Happy/Cheerful | ***0.61*** | 0.30 | 0.05 |
| Q4: Irritable | -0.06 | ***0.87*** | -0.04 |
| Q1: Anxious | -0.04 | ***0.86*** | -0.10 |
| Q2: Manage Behaviors | 0.14 | ***0.58*** | 0.24 |
| Q7: Sleep | -0.11 | 0.03 | ***0.93*** |
| Q6: Eat | 0.42 | -0.13 | ***0.40*** |

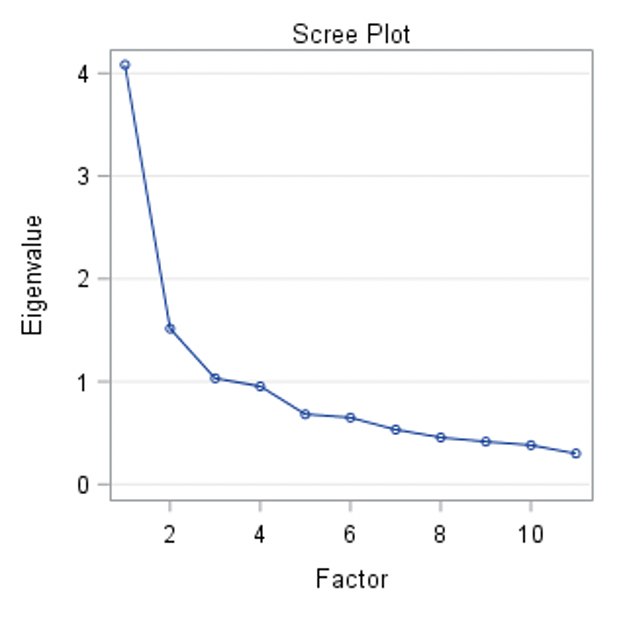

**Figure S2.** Scree plot examination of the behavioral survey factor structure.
